# Community access to rectal artesunate for malaria (CARAMAL): a large-scale observational implementation study in the Democratic Republic of the Congo, Nigeria and Uganda

**DOI:** 10.1101/2021.12.10.21266567

**Authors:** Christian Lengeler, Christian Burri, Phyllis Awor, Prosciova Athieno, Joseph Kimera, Gloria Tumukunde, Irene Angiro, Antoinette Tshefu, Jean Okitawutshu, Jean-Claude Kalenga, Elizabeth Omoluabi, Babatunde Akano, Kazeem Ayodeji, Charles Okon, Ocheche Yusuf, Nina C Brunner, Giulia Delvento, Tristan Lee, Mark Lambiris, Theodoor Visser, Harriet G Napier, Justin M Cohen, Valentina Buj, Aita Signorell, Manuel W Hetzel, the CARAMAL Consortium

**Affiliations:** Swiss Tropical and Public Health Institute, Basel, Switzerland; University of Basel, Basel, Switzerland; Makerere University School of Public Health, Kampala, Uganda; Kinshasa School of Public Health, Kinshasa, Democratic Republic of the Congo; Akena Associates, Abuja, Nigeria; Clinton Health Access Initiative, Boston, MA, USA; UNICEF, New York, NY, USA

**Author notes:** have equally contributed. Order in first pair drawn randomly. All additional members of the CARAMAL Consortium listed under Supporting information S1 Table.

**Keywords:** Malaria, pre-referral treatment, rectal artesunate, artesunate suppositories

## Abstract

The key to reducing malaria deaths in highly endemic areas is prompt access to quality case management. Given that many severe cases occur at peripheral level, rectal artesunate (RAS) in the form of suppositories was developed in the 1990s. RAS allows the rapid initiation of life-saving antimalarial treatment, before referral to a health facility with full case management capabilities. One randomized controlled trial published in 2009 showed a protective effect of RAS pre-referral treatment against overall mortality of 26%, but with significant differences according to study sites and length of referral. Two important issues remained unaddressed to-date: (1) whether the mortality impact of RAS observed under controlled trial conditions could be replicated under real-world circumstances; and (2) clear operational guidance for the wide-scale implementation of RAS, including essential health system determinants for optimal impact.

From 2018 to 2020, the Community Access to Rectal Artesunate for Malaria (CARAMAL) project was conducted as a large-scale observational implementation study in Nigeria, Uganda and the Democratic Republic of the Congo (DRC). CARAMAL aimed to provide high-quality field evidence on the two issues above, in three remote settings with high malaria endemicity. In order to achieve this, a number of complementary study components were implemented. The core of the CARAMAL study was the Patient Surveillance System (PSS), which allowed to track cases of severe febrile illness from first contact at the periphery to a referral health facility, and then on to a Day 28 visit at the home of the patient. Community and provider cross-sectional surveys complemented the PSS.

Here we describe in some detail RAS implementation, as well as the key CARAMAL study components and basic implementation experience. This manuscript provides an extensive reference document for the companion papers describing the impact, referral process, post-referral treatment and cost-effectiveness of the RAS intervention.

## Introduction

### Malaria globally

Malaria is one of the leading causes of illness, death, and lost economic productivity globally. Malaria case incidence per 1,000 population at risk decreased from 80 in 2000 to 57 in 2019, but annual reductions have significantly plateaued since 2015 [1]. Over the same period, malaria deaths decreased from 736,000 to 409,000 and this number has also stagnated since 2015. The heaviest malaria burden still affects sub-Saharan African countries that accounted for an estimated 90% of malaria cases and 92% of malaria deaths in 2019. Children under 5 years of age represented 84% of malaria deaths in 2000 and 67% in 2019 [1], and the risk of malaria is disproportionately high among hard-to-reach populations [2].

### Severe malaria

Mortality from malaria is due to uncomplicated disease from a Plasmodium infection left untreated and progressing to severe disease and ultimately death [3, 4]. Mortality from untreated severe malaria is very high, and varies according to epidemiological, biological and clinical severity factors [5–7]. Our best estimate of case fatality ratios (CFR) for African children with confirmed severe malaria disease and optimal treatment comes from the multi-centre AQUAMAT study [8]. Under parenteral artesunate treatment, CFR among 230 patients was 8.5% at 28 days following admission. On the other hand, CFR under routine treatment situations varies widely below or above this value because of varying quality of care, more or less stringent severe malaria case definitions, and lack of reliable cause-of-death estimates [6]. Effective clinical management of severe malaria patients has a high potential for reducing case fatality from malaria, with an estimated protective efficacy for routinely admitted children ranging from 76-87% [5]. In addition, effective treatment can prevent sequelae including developmental impairment [9], the recrudescence of infections, and ultimately also the emergence and spread of resistant parasites [10].

Reducing child mortality from malaria is arguably the first priority of any malaria control programme in highly endemic areas. To document possible reductions, national-level all-cause under-five mortality estimates are widely available thanks to Demographic and Health Surveys (DHS) and Malaria Indicator Surveys (MIS) carried out every few years. However, the causes of death, where and when these thousands of deaths occur, as well as the underlying social, economic and health system circumstances are not well documented. Few representative studies have investigated these on a large scale, as for example de Savigny *et al.* [11] and Gomes *et al.* [12]. Uncertainties remain with regard to the community rates of severe disease and the frequency of the different clinical presentations in different epidemiological settings [6]. In addition, access to and quality of care varies greatly between settings, with potentially significant impact on health outcomes [13].

Death from severe malaria often occurs within hours of signs of severity appearing. It is thus essential that therapeutic concentrations of a highly effective antimalarial drug be achieved as soon as possible, and a full treatment according to WHO guidelines be completed [10]. Synthetic derivatives of artemisinins are safe and fast acting, and ideally suited for treating severe malaria parenterally [8, 14, 15]. While the management of severe malaria cases is usually within the stated capabilities of all higher-level health facilities, in practice many practical issues such as chronic stock-outs, insufficient infrastructure and equipment, and inadequate human resources prevent the provision of quality care [13]. Access, in a broad sense, has been identified as a key obstacle to malaria treatment and care [16]. The current stagnation in rethe number of deaths from malaria has a various causes, but undoubtedly lack of access to timely high-quality malaria case management is a major one.

### Rectal artesunate (RAS)

As early as 1995 [17] it was shown that a single dose of artesunate, given rectally to adolescents and adults over 15 years, can achieve parasiticidal blood concentrations within 10–20 min, and can halve parasite density within 6–12 h [18–21]. Artesunate suppositories are easy to administer, safe, well accepted and allow rapid initiation of effective malaria treatment in remote locations [21].

However, one pre-referral dose of artesunate is nowhere near a complete treatment for severe malaria, and it is imperative that the full recommended curative treatment [10] follows the initial rectal artesunate (RAS) administration. The full recommended treatment sequence for severe malaria consists of intravenous or intramuscular artesunate for at least 24 hours, continuing until the patient can tolerate oral medication. Each patient should then receive a full course of artemisinin-based combination therapy (ACT) over 3 days [10]. The latter is particularly important to ensure full parasite clearance and to avoid artesunate monotherapy.

Following the first studies on artemisinin suppositories in 1995-1996, the Special Programme for Research and Training in Tropical Diseases (TDR) recognized in 1996 the need for a non-oral pre-referral treatment for severe malaria. In two clinical trials in 1996/1997, 50 mg and 200 mg capsules provided by Mepha Pharma of Switzerland were assessed [19]. These studies showed that the available doses were suboptimal for administration to children younger than 6 years, and a 100 mg dosage was suggested as an alternative (https://www.severemalaria.org/resources/rectal-artesunate-landscaping-assessment-report, accessed 15 November 2021). None of these earlier formulations were ever approved by regulatory authorities. Despite these shortcomings, the 50 mg and 200 mg capsules were added to the WHO Essential Medicine List in 2007 [21] and procured by countries, donors and non-governmental organisation (NGOs) until 2014, when a multi-donor-agency taskforce restricted donors from sourcing any anti-malarial products that were not approved by a stringent regulatory authority. Following a concerted action led by the Medicines for Malaria Venture (MMV) product development partnership, two RAS products (made by Cipla Ltd. and Strides Pharma Science Ltd.) became WHO-prequalified in 2018. Subsequently, they were registered in a large number of endemic countries and became widely available. In the same year, in preparation for large-scale deployment of RAS, the WHO issued revised guidance for RAS use as pre-referral treatment [22].

Unfortunately, local policies and practical guidance on the implementation of RAS are often not fully aligned with the WHO recommendation. A 2018 review of the national malaria treatment guidelines of all 56 African countries undertaken as part of the CARAMAL project found that just over half of the countries surveyed (n = 29) had included RAS in their national malaria treatment guidelines (https://www.severemalaria.org/resources/rectal-artesunate-landscaping-assessment-report, accessed 15 November 2021). Only 13 of these 29 countries had recommendations aligned with the WHO recommendation of treating only children under the age of six years with RAS, with five of these 13 countries recommending RAS for age five and below. The remaining 16 countries included the use of RAS in adults (or cited unclear age limitations), which is not recommended by WHO at present [22].

### Public health impact of RAS

To-date, 14 randomized controlled clinical trials have documented the safety, tolerability, efficacy and the pharmacological characteristics of RAS [21]. But only one large-scale Phase 3 field trial conducted in 2000-2009 assessed the public health impact of introducing RAS as pre-referral treatment in two African settings (Ghana and Tanzania, children between 6-72 months) and in Bangladesh (all ages) [12]. 17,826 patients suspected to suffer from malaria and unable to take oral medication were randomized to either RAS pre-referral treatment or to placebo, of which 12,068 were included in the final analysis. Suppositories were administered by local village residents with little medical training. After the administration of RAS, patients were referred to the nearest health facility offering parenteral treatment. In this study, 87% of patients in the African sites successfully arrived at the referral health facility, a very high percentage facilitated in part by the fact that transport was provided in some cases. That trial showed an overall protective effect against mortality of 26%, but with a high site-related heterogeneity (Africa versus Asia), and differences related to the length of time taken to reach a referral clinic. A significant protective effect was not seen for African children reporting to a referral clinic within less than 6 hours: 2.6% death/disability in the artesunate group, 3.3% in the placebo group (21.2% protective efficacy, not statistically significant at the 5% level). However, a statistically significant protective effect was seen for children alive after 6 hours, but not yet at a referral facility: 2.0% death/disability in the artesunate group, 3.9% in the placebo group, 48.7% protective efficacy, p<0.05. Unexpectedly, the study found increased mortality in older children and adults in Bangladesh, which was considered to be a chance finding in a 2014 Cochrane review [23].

In an accompanying commentary to [12], von Seidlein [24] asked the question of whether these results, obtained in the frame of a randomized clinical trial, were replicable in real-life settings. Given the importance of reducing deaths from malaria in highly malaria-endemic areas, and given our current lack of understanding of rates and determinants of fatal malaria disease, this question needed urgently to be addressed. The question re-surfaced at the time of the development of the new formulations of RAS around 2016, and triggered the launching of the “Community Access to Rectal Artesunate for Malaria (CARAMAL)” project in 2017.

### The CARAMAL project

The CARAMAL project, funded by UNITAID, was designed to support the market introduction of the two WHO quality-assured RAS products from Cipla Ltd. and Strides Pharma Science Ltd. CARAMAL aimed primarily at advancing the development of operational guidance for the implementation and scale-up of RAS. To do this, it relied on two essential and independent components: Firstly, RAS implementation in the frame of established integrated Community Case Management (iCCM) programmes, or in the frame of the Integrated Management of Childhood iIlnesses (IMCM) in low-level primary health facilities. Secondly, a large operational research component accompanying the implementation of RAS. The research component of the CARAMAL project tested the hypothesis that it is feasible to achieve reductions in severe malaria case fatality ratios by delivering RAS through established routine health system platforms, without substantial supportive interventions, and without unintended negative consequences.

Here we present a general description of the CARAMAL project, its research questions, detailed methodology and overall operational results. The key findings and recommendations from the scientific investigations of the CARAMAL project are presented elsewhere, with a focus on the health impact of the large-scale roll-out of RAS [25], referral processes and completion rates [26], the quality of treatment at referral health facilities [Signorell *et al.* manuscript in preparation], and cost-effectiveness [Lambiris *et al.* submitted]. Given its size (over 14,000 episodes of severe fever observed), its ability to follow these children for 28 days after enrolment into the study, and the multiplicity of research tools, CARAMAL offered unique insight into the fate of children experiencing severe febrile illnesses (including severe malaria) in remote locations in Africa. The study also illuminated the many health system characteristics that led either to successful or to sub-standard antimalarial treatment effectiveness, ultimately determining child survival.

## Materials and Methods

### Project setup

The CARAMAL project was organized as a large multi-country consortium. The Clinton Health Access Initiative (CHAI) provided overall management. The United Nations Children’s Fund (UNICEF), together with national and local health authorities, were responsible for introducing and distributing quality-assured RAS at primary health care level through health care providers that lack the capacity to administer parenteral treatment. This included community health workers in iCCM programs, as well as primary health care facilities. The operational research component was coordinated by the Swiss Tropical and Public Health Institute, and implemented in the Democratic Republic of the Congo (DRC) by the Kinshasa School of Public Health, University of Kinshasa; in Nigeria by Akena Associates Ltd.; and in Uganda by the School of Public Health of Makerere University. The Pasteur Institute in Cambodia was contracted to provide an assessment of molecular markers of artemisinin resistance in the study areas. Medicines for Malaria Venture (MMV) was responsible for making quality-controlled pre-qualified RAS available globally and in the three project countries. The WHO Global Malaria Program (WHO GMP) provided technical and scientific support on specific aspects of the project, and was responsible for turning the generated evidence into policy and implementation guidelines.

### Country and site selection

The CARAMAL project was implemented in three sub-Saharan African countries, each with a high malaria burden. The three project countries (DRC, Nigeria, and Uganda) together share about 42% of total global malaria cases (DRC 19 million cases, Nigeria 61 million cases, Uganda 8.5 million cases in 2019) and 39% of total reported malaria deaths (DRC 42,000 deaths, Nigeria 110,000, Uganda 12,000 in 2019)[1].

The study areas included three Health Zones in the Provinces of Kwilu and Kwango in DRC, three Local Government Areas (LGA) in Adamawa State in Nigeria, and three districts (Oyam, Kwania & Kole) in the Northern Region of Uganda. In total, the study areas covered a population of approximately 2.5 million, including 476,000 children under five years (Fig 1 and Supporting Information S1 Fig).

**Fig 1:**
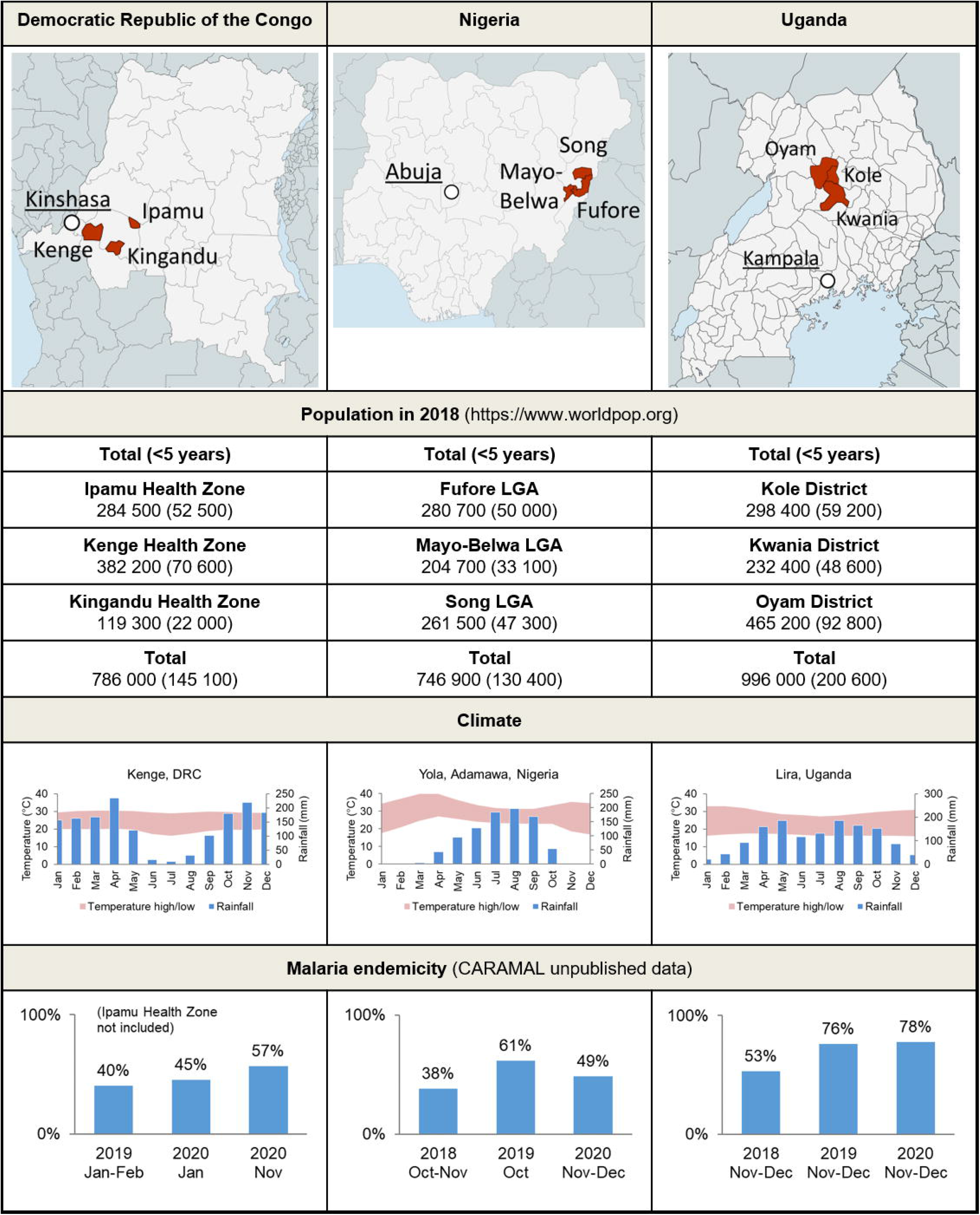
Study areas and study populations in the three CARAMAL project countries. mRDT = malaria Rapid Diagnostic Tests.

The study sites in the three countries were selected based on the evaluation by UNICEF - and vetted by national authorities - of ongoing iCCM operations, case numbers, and security considerations. All implementation sites were considered remote based on distance to tertiary health facilities, to reflect settings in which RAS could have the greatest potential based on previous findings [12]. To provide some degree of geographical, cultural, linguistic and health system diversity, one country was selected from West, Central and East Africa, respectively. Fig 1 also shows key climatic data, as well as malaria infection rates in children under five years of age, as measured in the frame of the CARAMAL study. The three sites were shown to have a high level of endemicity: the community parasite rates in children under 5 years were 40-57% in DRC, 38-61% in Nigeria and 53-78% in Uganda (Fig 1).

### Health system environment

Health services in the study sites consisted of a network of formal health facilities, including primary health centres (PHC) that offered mainly preventive and curative outpatient services, and referral health facilities with higher-level medical capabilities and usually inpatient wards (Table 1). In all three countries, and following national guidelines, comprehensive treatment for severe malaria patients was only meant to be provided in referral health facilities. Private for-profit clinics and pharmacies were present in each site in varying numbers. Unfortunately, we could not cover these fully in the frame of the CARAMAL work because this would have exceeded the available resources. But patient visits to these facilities were elicited in the various data collection instruments.

**Table 1:** Public health care providers, estimated under-five population, and number of under-fives per provider in the study areas, by provider type and country (all data for 2018).

|  | **DR Congo** | **Nigeria** | **Uganda** |
| --- | --- | --- | --- |
| Community Health Workers | *Sites de Soins Communautaires* (Community Care Sites)<br>N = 42 | Community-Oriented Resource Persons (CORP)<br>N = 500 | Village Health Team (VHT) members<br>N = 5,100 |
| Primary Health Care facilities | *Postes de Santé* (Health Posts)<br>*Centres de Santé* (Health Centres)<br>N = 152 | Health Posts<br>Primary Health Centres (PHC)<br>N = 77 | Health Centres II (HC II)<br>N = 30 |
| Referral Health Facilities | *Centres de Santé de Référence* (Referral Health Centres) and<br>*Hôpitaux Généraux de Référence* (General Referral Hospitals)<br>N = 19 | Cottage Hospitals<br>N = 3 | Health Centres III (HC III),<br>Health Centres IV (HC IV)<br>and Hospitals<br>N = 20 |
| Number of U5 children | 145'107 | 130'430 | 200'518 |
| U5 per community-based provider | 690 | 284 | 46 |
| U5 per RHF | 7,045 | 55,022 | 11'816 |

RAS was implemented in all Primary Health Care (PHC) facilities. RAS was also implemented in each study site through Community Health Worker (CHW) programmes offering basic preventative and curative services, in line with global and national iCCM guidance. The CHWs implemented iCCM algorithms, which could be leveraged in the frame of the CARAMAL project to implement RAS [27]. In this manuscript, community-based providers in all three countries are called “CHW”, even though there was a different terminology in each country. Their numbers also differed significantly by country (Table 1). In all three countries, CHWs were unpaid volunteers or individuals receiving only a minimal financial incentive. The number of health care providers, the estimated under 5 population, and the ratio of under-fives to healthcare worker in the three CARAMAL project sites are all shown in

**Table 1**. Uganda had both the highest coverage of referral health facilities (RHFs) and community-based providers (CHW and PHC). In DRC and Nigeria, CHWs are strategically located in locations where other formal public health providers are far away (i.e. considered hard-to-reach). In Uganda, the national policy is that two CHWs are located in every village, explaining their high coverage. Table 1 illustrates the significant differences in the health care structure between the three sites.

### Main intervention: rectal artesunate (suppositories)

WHO-prequalified and in-country registered RAS suppositories each containing 100 mg of artesunate (manufactured by Strides Pharma Sciences Ltd. or Cipla Ltd., both in India), were purchased and shipped to the project countries by UNICEF. The manufacturer’s dosage recommendations were one suppository of 100 mg for children between 6 months and 3 years, and two suppositories for children from 3 to 6 years of age. In the three countries, RAS pre-referral treatment is part of the countries’ current national malaria treatment policy, though not yet implemented at scale. In the study areas, RAS was rolled out at community level in early 2019 through the two channels list above.

In preparation for the roll-out of RAS, MMV developed job aids and training materials for the appropriate management of children experiencing symptoms of severe febrile illness/suspected severe malaria (Fig 2). According to WHO/UNICEF iCCM guidelines [27], patients eligible for pre-referral RAS are defined as follows: age below 5 years, fever or a history of recent fever, plus one of the following general danger signs : lethargy/unconsciousness, not able to feed or drink, convulsions, repeated vomiting. While local iCCM guidelines (e.g. in Nigeria) included a few additional danger signs, it was this broad clinical picture of severe febrile illness – especially the inability to tolerate oral medications, that prompted the administration of RAS and the issuing of a referral notice to a higher level health facility. Of note is that (1) the iCCM definition of a severe febrile illness does not overlap entirely with the clinical definitions of severe malaria [10], and (2) the iCCM algorithm does not require a malaria Rapid Diagnostic Test (mRDT) if severe malaria is suspected. These points will be further elaborated in the results section below. Patients attending primary health centres staffed with medical personnel were usually assessed based on the IMCI criteria, and administered RAS based on a severe malaria diagnosis.

**Fig 2:**
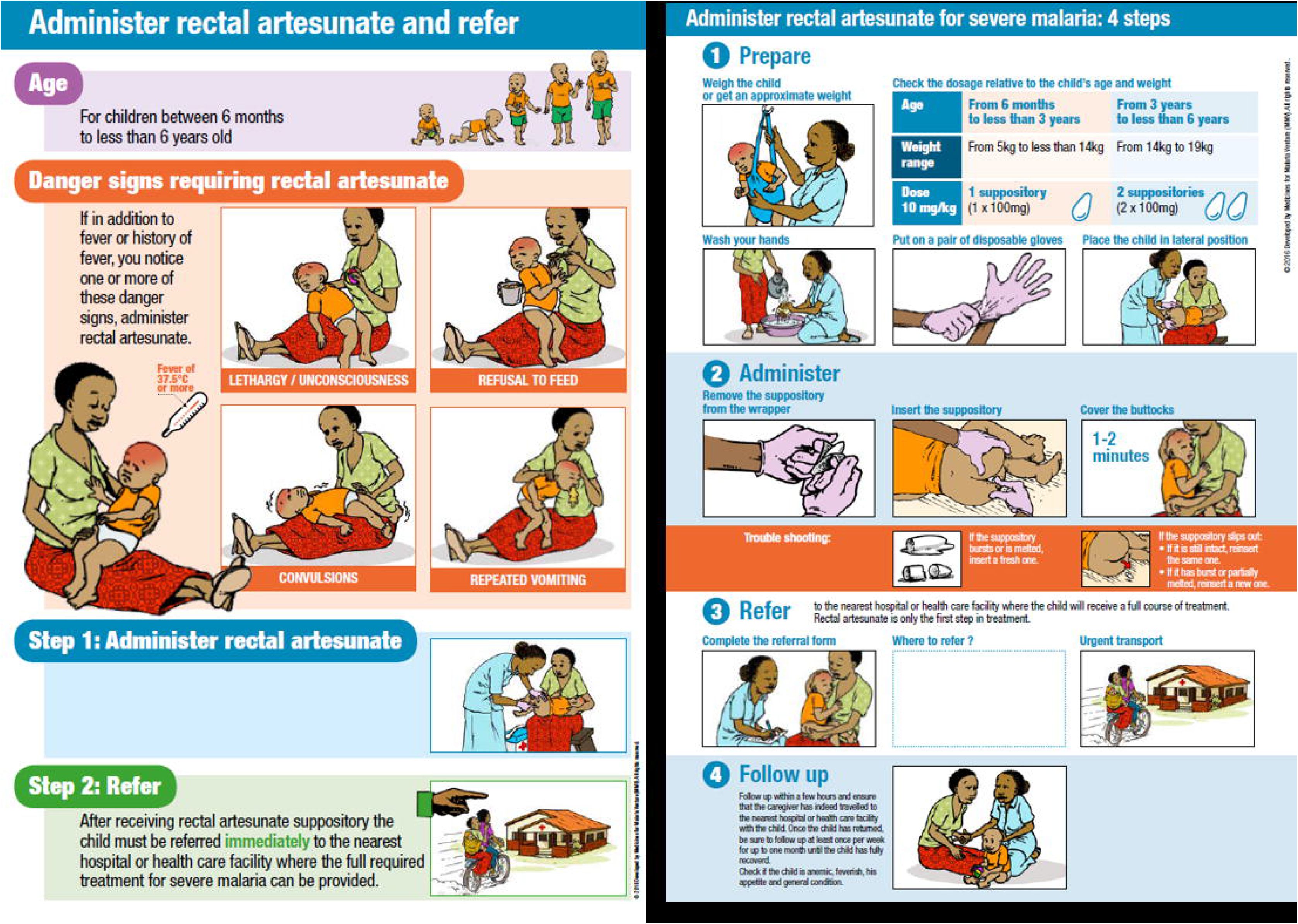
Teaching aids developed by Medicines for Malaria Venture (MMV) for the training of health workers in the correct administration of RAS (source: https://www.mmv.org/access/tool-kits/artesunate-rectal-capsules-tool-kit).

Training sessions on RAS administration were conducted for over 8,000 CHWs and staff of primary health facilities in 2019 - in some instances embedded in broader training/retraining of CHWs. A second round of trainings was conducted in mid-2020 for new staff and as a refresher for previously trained staff. These were also adapted to include content on infection prevention and control in the context of the COVID-19 pandemic.

### Supportive health system interventions

A limited number of essential supportive interventions were implemented in addition to introducing RAS, to facilitate rollout and foster appropriate higher-level care in the study sites. In DRC, additional injectable artesunate had to be distributed to referral facilities (27,000 vials in 2019) even though this was meant to have already been included in ongoing malaria control programmes. In Nigeria, a color-coded, two-way referral form was printed to improve communication between CHWs and referral health facility staff; and an existing emergency transport system for emergency obstetric care was extended to allow the transport of severely ill children. In Uganda, district-level (parish) coordinators were engaged to support data collection from CHWs and support replenishment of their RAS stock. These additional health system strengthening activities were limited to a few essential supplies and processes by design due to scalability considerations, and purposely did neither include large-scale supply chain management support nor fundamental improvement of referral services given the aim of evaluating the effect of introducing RAS under real-world circumstances. The issue of transport subsidies to facilitate referral completion was discussed within the team, with relevant national authorities and the donor. For the same considerations as above it was deemed that additional health systems interventions such as transport subsidies or purchase of additional transport modalities (e.g. motorcycles), while potentially life-saving, would be unsustainable in the long-run and thus outside the aim of generalizability of the CARAMAL project.

A round of supportive supervision was initiated shortly after RAS rollout in April 2019 by the relevant government authorities to ensure proper clinical care and effective utilization of supplies. While supportive supervision and review meetings are considered a key part of community-level primary health care (as they are used to train new CHWs and peripheral health facility staff, ensure quality of care, collect new supplies, return expired RAS, submit reports, etc.), regularity is often impeded by a number of factors including lack of financing, transport, climatic & security factors. Project implementation was concluded in October 2020 and activities were transitioned to national authorities in each project country by April 2021.

### Study design and conceptual framework

Evidence generation around the roll-out of RAS was designed as an observational study with a before-and-after plausibility evaluation design [28]. Given (1) the massive sample size required (total population exceeding 2,200,000, including 400,000 children under five years - see calculations below), (2) the enormous practical implementation difficulties due to the remote location of the study sites, and (3) ethical issues in running a control group for an established life-saving intervention already included in national treatment guidelines, this was de facto the only possible design. We are well aware that such a before-and-after design bears a high risk of bias for the main outcome – impact on case-fatality – because many underlying contributing factors (seasonal and climatic, health system functioning, economic, co-interventions for malaria prevention, disease outbreaks etc.) could have an uncontrolled impact on the outcome measurements. To mitigate this problem, as many of these factors as possible were measured through the different data collection tools (see sections below), triangulated, and key co-variates were used in the multivariable analyses that are presented in forthcoming publications.

The overall conceptual framework for evidence generation was the continuum of care for severe febrile illness: starting with the recognition of a child’s illness at home, then care-seeking and case management outside the home, and ending 28 days later with the determination of the health status of the child (Fig 3). This continuum was broken down into discrete ‘steps’ that could be measured as part of a real-world effectiveness study.

**Fig 3:**
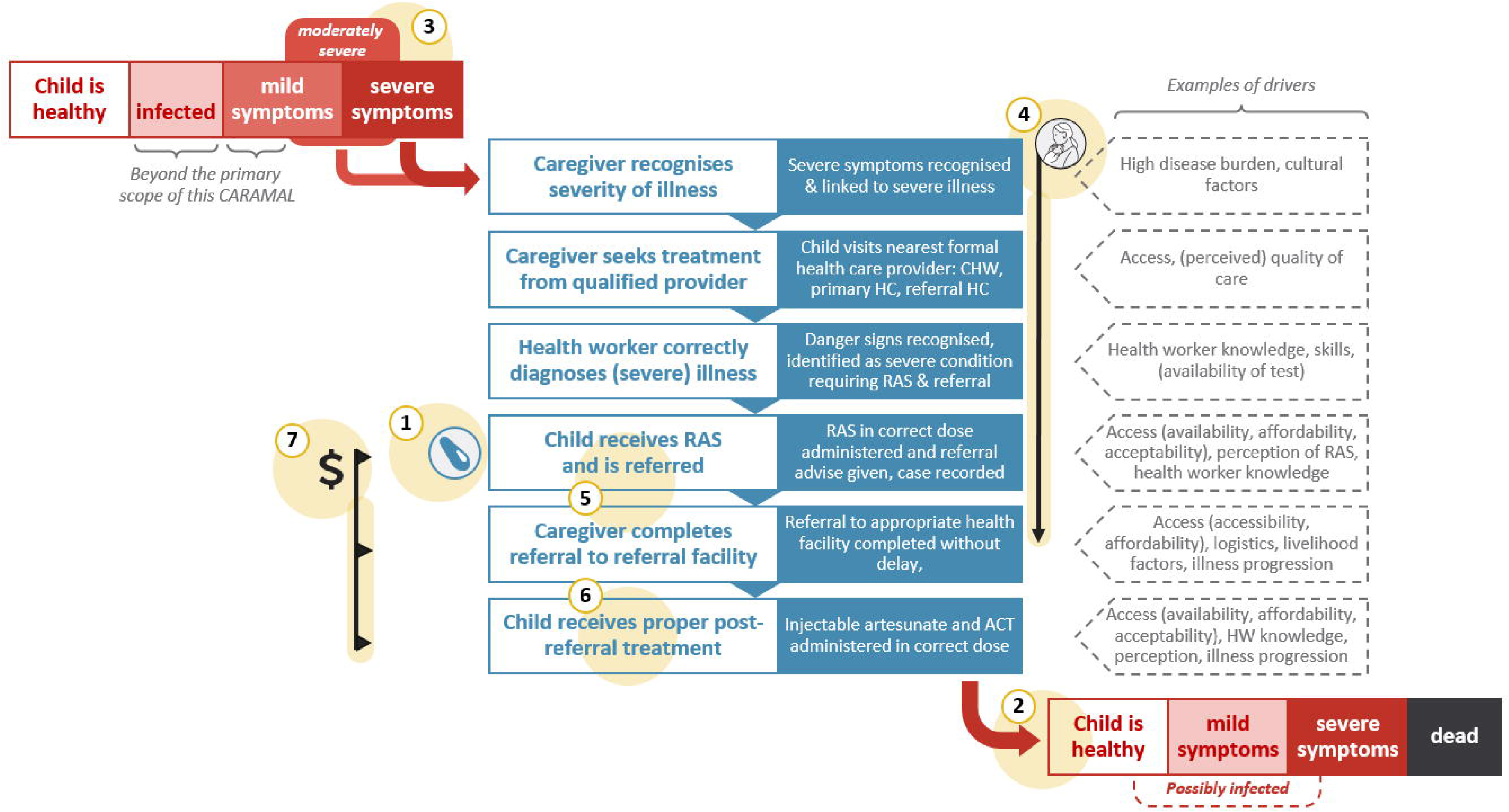
Continuum of care for an episode of severe febrile illness (central block) and key themes identified for analysis (numbered with yellow highlights). 1. RAS implementation (coverage), 2. Health impact of introducing pre-referral RAS, 3. Severity of illness, 4. Treatment seeking pathways, 5. Treatment and referral at community-based providers, 6. Case management at referral facilities, 7. Cost and cost-effectiveness of introducing RAS.

### Study population

The CARAMAL study components focused on children <5 years of age (rather than < 6 years as in the WHO guidelines for pre-referral RAS use [2]), to be in line with the target age group of iCCM and IMCI guidelines [27]. The Patient Surveillance System (PSS) described below covered primarily children seeking care for a current episode of severe febrile illness starting at the level of a community–based health care provider. This included suspected severe febrile illness patients attending a CHW and identified based on iCCM general danger signs, as well as patients attending a PHC and identified as cases of severe malaria.

In addition, children <5 years directly seeking care at a referral facility and admitted with a diagnosis of “severe malaria” were also enrolled. Even though these children were not eligible for RAS pre-referral treatment, and thus not part of the primary analysis, they were included in the CARAMAL study to understand their comparative treatment seeking behaviour, diagnosis, and case management at the referral facility. Including these patients also allowed for a comparison between case numbers and CFR at community-based providers versus referral facilities, and a more comprehensive calculation of severe febrile illness / severe malaria rates.

### Data collection methods

Data were collected through a set of complementary activities aligned with the roll-out of RAS in the study areas (Fig 4). RAS roll-out was gradually initiated from March-April 2019 onwards. Pre-RAS implementation baseline data was collected between April 2018 and March-April 2019, post-RAS implementation data between March-April 2019 and August 2020, with small local variations depending on the RAS roll-out progress.

**Fig 4:**
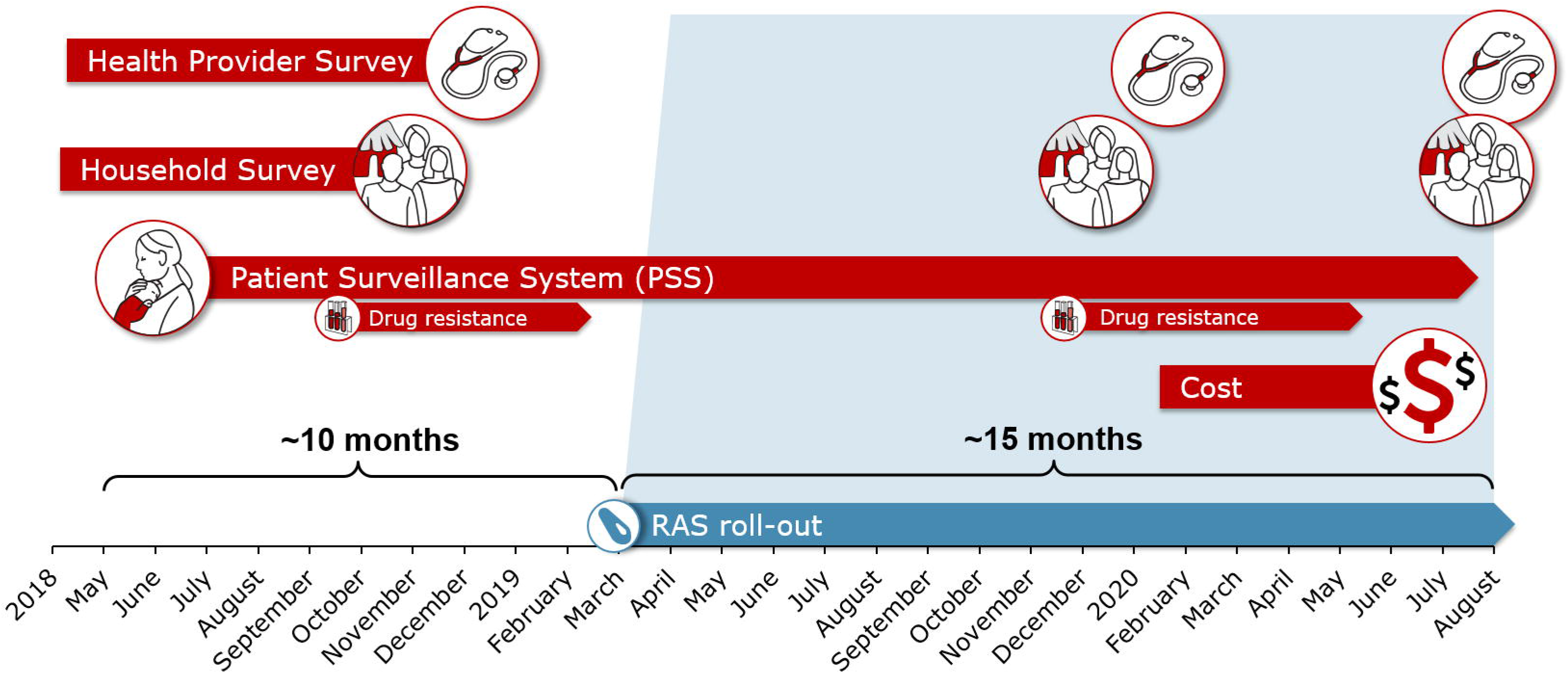
Schematic representation of the various CARAMAL survey instruments and the duration of data collection.

The core of the CARAMAL evidence generation was the PSS, described in detail below. Patients were enrolled in the PSS over the entire study period, both before and after the roll-out of RAS. Data on molecular markers of artemisinin drug resistance were collected in the frame of the PSS (see below). In addition, one health care provider survey (HCPS) and one household survey (HHS) took place during the pre-RAS period, and two each in the post-RAS implementation period. In DRC, the last HCPS and HHS were only conducted in 2 out of the 3 sites. Data for the costing study were collected in 2020. Data collection activities are described in detail below; a comprehensive list of indicators related to each activity is provided in <u>Supporting information S2 Table</u>.

### Activity 1: Patient Surveillance System (PSS)

Patients were tracked at three contact points during the health seeking pathway (Fig 5): (1) at the CHW or primary health care facilities, where patients were provisionally enrolled and assigned a unique study ID, (2) at the referral facilities, and (3) during a follow up visit at home 28 days after the first contact. Alternatively, for children found to be deceased by day 28, an adapted interview was conducted about one month after their passing to respect the mourning period.

**Fig 5:**
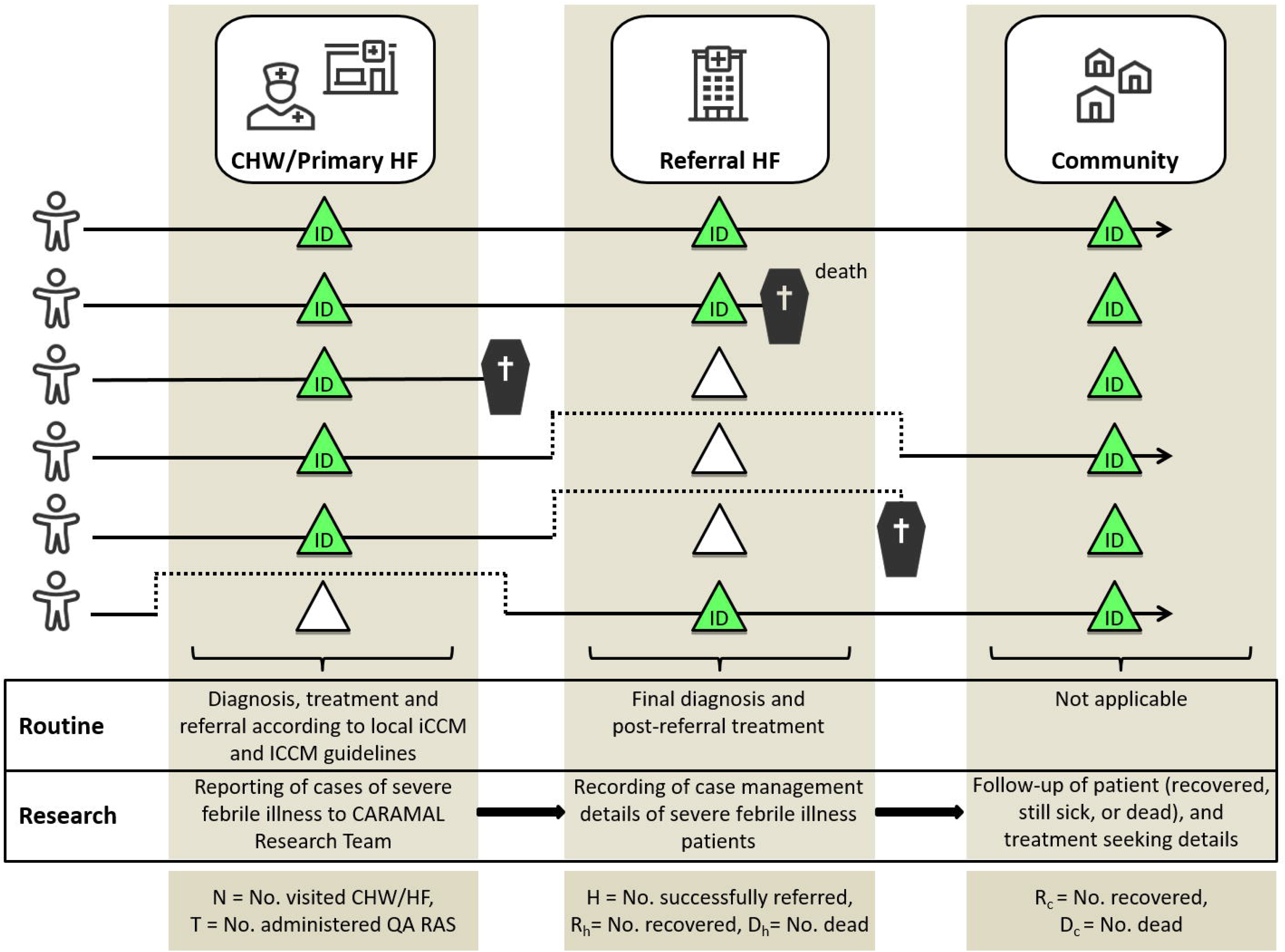
Schematic representation of possible points of contact in the Patient Surveillance System. ID = identification by unique study ID. Green triangles represent points of contact, red triangles absence of contact. CHW=Community Health Workers. HF=Health facility. No=Number. QA=Quality-assured.

At the CHW or primary health facilities, the children underwent standard clinical examinations, with treatment and referral procedures as per local iCCM or IMCI. For CARAMAL study purposes, a rapid diagnostic test for malaria (mRDT) was also conducted at the primary point of contact for all eligible children.

CARAMAL study nurses were posted at major referral facilities to document the triaging, registration procedures, diagnosis and treatment of patients admitted with severe malaria, either after referral from a community-based provider, or presenting directly to the referral facility. The study nurses also recorded the patients’ signs and symptoms, clinical progression, discharge outcome (recovery, death) alongside the approximate timing of all procedures. Data were captured in real-time, through observation or extracted from routine hospital records, as appropriate. All enrolled patients were reported to the study team and registered in the study database. On this basis, a follow-up home visit after 28 days was scheduled.

At the day 28 home visit, a structured questionnaire was used to elicit information on the current health status of the child (cured, still ill, passed: primary outcome) and care seeking since the time of first contact, with a special focus on the referral process and antimalarial treatment. Perception of RAS and care seeking costs were also elicited. An mRDT and haemoglobin (Hb) measurement (HemoCue Hb 201, Ängelholm, Sweden) were performed using capillary blood. Due to the high prevalence of positive mRDTs at the day 28 follow-up visits early in the project, as well as evidence of prolonged *Plasmodium falciparum* histidine rich protein 2 (HRP2) antigenaemia [29], a decision was taken in January 2019 to switch from an HRP2-only test to an HRP2/pLDH (Pf/PAN) combo test. *Plasmodium sp*. lactate dehydrogenase (pLDH) is eliminated much more rapidly after successful treatment [29], allowing for a more accurate estimate of the number of patients with an active infection on day 28.

The day 28 visit and the monitoring of patients during their admission at referral facilities were the primary data sources for the monitoring and evaluation of patients. In addition, selected data points were collected directly from the enrolling community-based providers, based on their routine records. This included crucially information on RAS administration. Data was collected in ODK Collect (https://opendatakit.org/) on tablet computers.

#### Sample size calculation for the PSS

Based on historical case-fatality reports for severe malaria (2.8% MATIAS Study DRC [30], 8.5% AQUAMAT [8]) a 6% CFR was assumed at baseline. The data collection time was 6 months pre-RAS (baseline) and 18 months post-RAS. The original plan to have one year baseline and two years post-RAS could not be realized after extensive delays due to the ethical approval process. The sample size calculation was based on the formulas by Fleiss 2003 [31] with an allocation ratio of 3, and computed in STATA using the ‘power two proportions p1 p2, nratio’ command.

A minimum sample size estimation of 6,032 cases of severe malaria in children <5 years over 24 months was based on the detection of a 30% decrease in case fatality across the three project countries following the roll-out of RAS, with 80% power and α = 0.05. Rounded up, this translated into 1,600 cases in the pre-RAS at baseline phase and 4,600 post-RAS roll-out, across the three countries (total 6,200). We also assumed that only 40% of all severe malaria cases would be enrolled into the PSS at CHW/primary health facility level and followed up on day 28. For measuring changes in CFR, a pooled analysis with data from the three countries was planned, while other less sample-size demanding indicators were calculated per country. In order to obtain this number of severe malaria cases, a total population of 2.2 million individuals including at least 400,000 children under 5 years needed to be considered. This target was exceeded (see Fig 1). Further calculations per country confirmed that this sample size was large enough that other conditions such as severe febrile illness, and improvements in referrals, could be detected with at least the same probability (calculations not shown).

### Activity 2: Health Care Provider Surveys (HCPS)

HCPS were conducted annually to assess the severe malaria case management capacity of the local health system. Two survey instruments were completed by each provider: (1) a structured checklist assessing the availability of essential medical supplies and equipment, human resources, infrastructure and documentation in their health facility, and (2) a questionnaire aimed at the providers themselves, and containing questions on demographics, education and training, work experience, supervision, training received, knowledge, attitudes and practices relevant to febrile case management (incl. diagnostic algorithm and RAS treatment guidelines), and implementation experience with these treatment guidelines. Both forms were designed for electronic data entry using ODK. All included health care providers signed an informed consent form before the interview.

#### Sampling and sample size calculation for the HCPS

The HCPS included a stratified random sample of health care providers treating children < 5 years, including 40 community-based providers (CHW) and staff at primary and referral health facilities. In DRC and Uganda, the survey included most non-referral higher health facilities in the study area, while in Nigeria, a random sample of 40 higher-level facilities was included. The following clinical/nursing staff involved in the treatment of children under five years with febrile illness were eligible for interviews: i) all selected CHW; ii) at selected primary health care facilities: the officers in charge and all other clinical staff; iii) at referral facilities: the officer-in-charge and all clinical staff in the out-patient and paediatric inpatient departments, among which two staff members per category were randomly selected for interview.

The sample size was estimated to allow detecting a 19% decrease in the minimum acceptable coverage (defined as availability of RAS and adherence to case management guidelines including referral), following the roll-out of RAS. From a baseline value of 80%, with a power of 80% and anα error = 0.05.

### Activity 3: Household surveys (HHS)

HHS were carried out annually to assess malaria control intervention coverage, caretaker treatment seeking behaviour, caretakers’ knowledge of and attitudes towards RAS, and to measure prevalence of malaria and anaemia in the population < 5 years of age (using the same procedure as during the Day 28 visit). In each household, the household head and parents/caregivers of children < 5 years of age were eligible to participate in face-to-face structured interviews, after informed consent. All forms were designed for electronic data entry using ODK. Sampling and sample size calculation for household surveys: Households were selected using a two-stage random sampling approach (village-household), whereas the sampling frames consisted of all villages in the study area and all households in the village with at least one child < 5 years. Treatment seeking from formal health facilities in case of fever in children <5 years was assumed to be between 15% [32] to over 75% [33]. For the calculation of the sample size an increase in treatment-seeking from 15% (minimum scenario pre-implementation) to 20% post-implementation of RAS, with 80% power and α = 0.05 was aimed for. A minimum of 906 household survey responses per country and survey round was required.

### Activity 4: Economic evaluation

Based on a combination of programmatic records and research data, this study component assessed the incremental economic costs of RAS introduction. In a second step, the incremental cost-effectiveness of RAS introduction over the current standard of care for the management of severe febrile illness / malaria was assessed. The methodology for this component is described in detail in a forthcoming publication (Lambiris *et al.*, submitted). Of note, funding from the CARAMAL project was always additional to funding provided by government resources and support by other partners.

Programmatic records of RAS implementation by UNICEF and its partners provided the backbone for the routine monitoring of process indicators. These included procurement records, supervision reports, routine reports and provider geo-locations, which were on occasion incomplete for a variety of factors.

### Effects of the COVID-19 Pandemic

Due to the COVID-19 pandemic and the resulting travel restrictions and government safety procedures, certain adaptations in data collection became necessary in 2020. Some Day 28 interviews were conducted intermittently by phone when no visits were possible. The last round of the Health Care Provider and Household Surveys was postponed beyond the planned period of data collection (September-October 2020) to December 2020 in DRC, when the areas were again fully accessible. Fortunately, the overall effect of the pandemic on data collection was not too high, even though some temporary service disruptions were noted and in all three countries.

### Monitoring of Artemisinin resistance

While resistance of the *Plasmodium* parasites against artemisinin derivatives had not yet been documented in the study settings in 2017, this prospect was of great concern to WHO and public health practitioners. Of special concern in the frame of the CARAMAL project was the fact that RAS is a monotherapy, and that the widespread use of artemisinin monotherapies could increase drug pressure. The same problem is obviously also arising from parenteral use of artesunate, the currently recommended drug of choice for severe malaria, if not followed by an oral ACT as per WHO recommendations [10]. According to market data provided by MMV (www.severemalaria.org/severe-malaria-market-situation), the expected annual market for RAS is one million units per year, against 25 million doses of injectable artesunate. Hence the issue of monotherapy is potentially more related to the uncontrolled use of injectables rather than RAS.

To monitor prospectively the frequency of resistance markers in the three study settings, a complementary study was implemented in collaboration with the WHO Global Malaria Programme (GMP). The project sampled children of different groups with malaria in the frame of the PSS at two time points (see Fig 4 above) to measure the prevalence of molecular markers of artemisinin resistance (K13-propeller sequence polymorphisms). Samples were collected as dried blood spots (DBS) on filter papers. The DBS were then sent to the WHO-contracted laboratory at the Institut Pasteur du Cambodge in Phnom Pen for K13 genotyping. Details of the methods and the results are presented in a forthcoming publication (Awor *et al.* manuscript in preparation).

### Data management

Quantitative data was collected by trained field research teams and study nurses at referral health facilities using tablet computers and the ODK Collect application. ODK forms were designed by the CARAMAL research team. Data were transferred from tablets to the study database (ODK Aggregate server) whenever a reliable internet connection was available. The data were stored on dedicated secured ODK Aggregate server partitions at the Swiss TPH in Basel, Switzerland, and access restricted to study staff directly involved in data management and analysis. An automated daily backup of all the study data was made. Where electronic data capture was not feasible, qualitative and quantitative data were captured on paper forms (or audio recorded if applicable), and then transcribed to ODK Collect. Regular spot-checks and data quality audits were carried out to assess completeness and coherence.

### Ethical considerations

The CARAMAL study protocol was approved by the Research Ethics Review Committee of the World Health Organization (WHO ERC, No. ERC.0003008), the Ethics Committee of the University of Kinshasa School of Public Health (No. 012/2018), the Health Research Ethics Committee of the Adamawa State Ministry of Health (S/MoH/1131/I), the National Health Research Ethics Committee of Nigeria (NHREC/01/01/2007-05/05/2018), the Higher Degrees, Research and Ethics Committee of the Makerere University School of Public Health (No. 548), the Uganda National Council for Science and Technology (UNCST, No. SS 4534), and the Scientific and Ethical Review Committee of CHAI (Date 21 Nov 2017, #112). In addition, the study protocol was accepted by the national and local health authorities in all three implementation countries. The study was registered on ClinicalTrials.gov (NCT03568344).

Informed consent from the participant’s parent or guardian was obtained prior to the inclusion into the PSS. Preliminary oral consent was asked at the primary care level because of the urgency of the child’s clinical condition. This oral consent was followed by a written consent either at the referral health facility, or during the day 28 follow-up at home, whatever came first.

Written consent was obtained separately from health workers prior to participating in the HCPS (Activity 2), and from household heads and parents/guardians of children < 5 years prior to participating in the HHS (Activity 3).

## Results and discussion

### Patient Surveillance System (PSS): key enrolment figures

Across the three countries, health care workers at first contact provisionally enrolled 14,911 patients into the PSS. Of these, 13,758 (92%) were fully enrolled, i.e. written informed consent was provided and the day 28 follow-up was successfully completed. Despite a comparable population of children <5 years in the three countries (130,000-200,000), there was a large difference in total enrolment numbers: DRC 5,540, Nigeria 1,505, Uganda 6,713.

Almost half of the children were enrolled directly at a referral health facility (6,131, 45%) (**Table 2**), while CHWs enrolled 4,415 (55%) and PHCs contributed 3,212 (23%) enrolments. In Nigeria, most enrolments came from RHFs. In DRC, a minority of enrolments came from CHWs (a reflection of the small number of these providers), while in Uganda, more enrolments came from CHWs than from PHCs, probably as a result of their large number (see also Table 1). Particularly in Nigeria, the proportion of PHC enrolments increased from pre-RAS (11%) to the post-RAS period (26%), but overall more children were recruited directly at the RHF level.

**Table 2:** Number of Patient Surveillance System (PSS) enrolments, by enrolment location, RAS implementation period, and country.

|  | DR CONGO |  | NIGERIA |  | UGANDA |  | TOTAL |
| --- | --- | --- | --- | --- | --- | --- | --- |
|  | Pre-RAS<br>N (%) | Post-RAS<br>N (%) | Pre-RAS<br>N (%) | Post-RAS<br>N (%) | Pre-RAS<br>N (%) | Post-RAS<br>N (%) | N (%) |
| CHW | 60 (4) | 104 (3) | 178 (30) | 180 (20) | 1608 (72) | 2285 (51) | 4415 (32) |
| PHC | 701 (40) | 2177 (57) | 67 (11) | 233 (25) | 1 (0) | 33 (1) | 3212 (23) |
| RHF | 973 (56) | 1525 (40) | 351 (59) | 496 (55) | 620 (28) | 2166 (48) | 6131 (45) |
| Total<br>(study<br>period) | 1734 (100) | 3806 (100) | 596 (100) | 909 (100) | 2229 (100) | 4484 (100) | 13'758 (100) |
| Overall<br>total | 5540 |  | 1505 |  | 6713 |  | 13'758 |

Details on place of recruitment, age and sex-distribution of the enrolled children are given in the <u>Supporting information S3 Table</u>. Enrolments by month and country are shown in Figs 6 a-c below.

**Fig 6:**
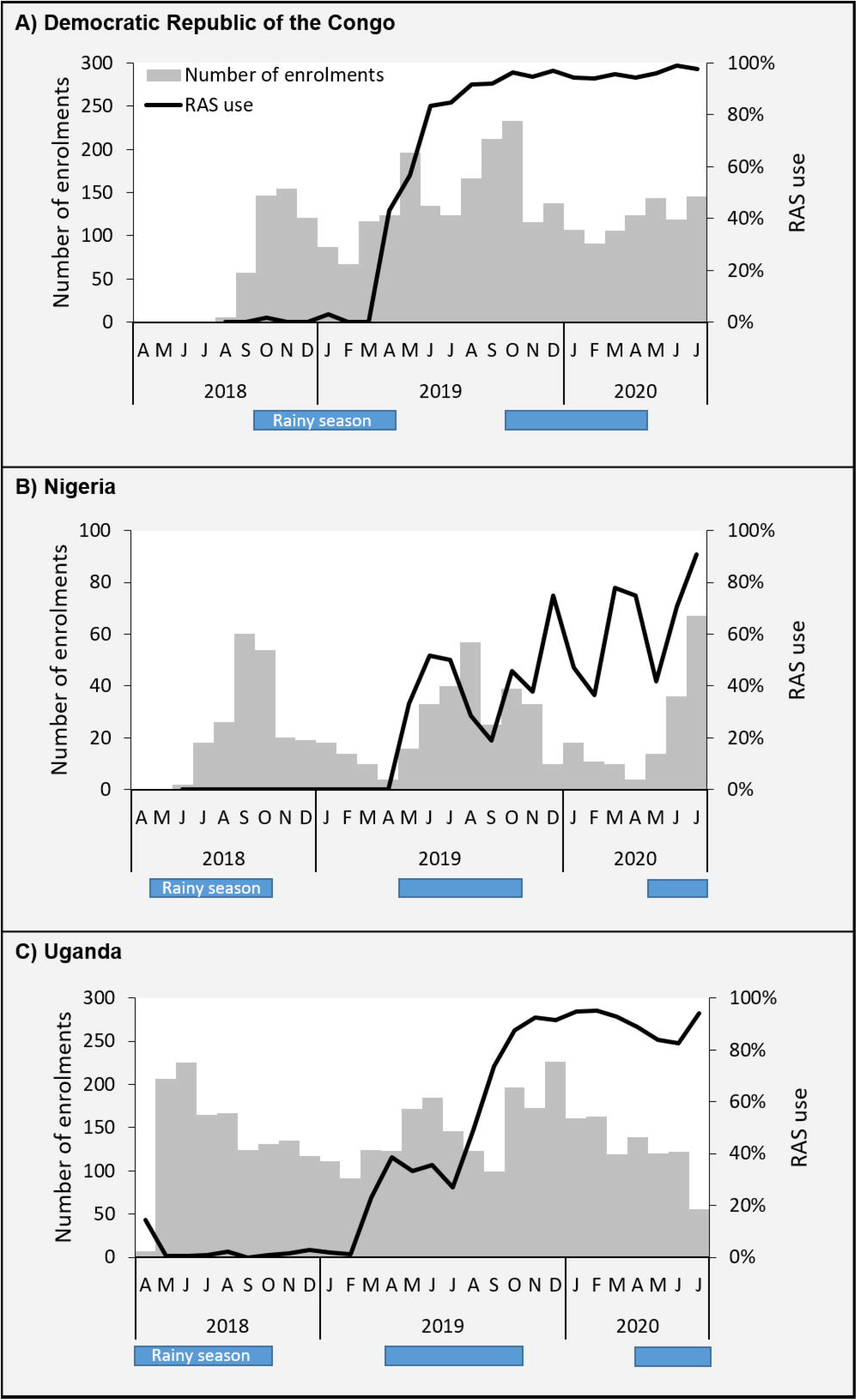
Number of children enrolled in the Patient Surveillance System (grey bars), and percentage of these children being administered rectal artesunate (RAS), by country.

### RAS rollout

Detailed information on operational aspects of RAS rollout are available in Lambiris *et al.* (submitted). Some key results are reported below. In DRC, the 100 mg RAS products from Cipla^TM^ and Strides Pharma Science^TM^ received marketing authorization in June 2017 and September 2017, respectively. RAS coverage increased rapidly and remained high after its initial introduction in March 2019 (Fig 6a). In Nigeria, the 100 mg RAS product from Strides Pharma Science^TM^ received marketing authorization in January 2019, and the product from Cipla^TM^ in October 2019. Two procurements were initially anticipated to cover the needs of the study areas through the end of implementation in October 2020. However, civil protest and riots in July-August 2020 resulted in the looting of RAS storage facilities in Adamawa. A total of 7,577 RAS units were lost in the attack. RAS coverage increased inconsistently after introduction in April 2019 (Fig 6b). In Uganda, both 100 mg RAS products from Cipla^TM^ and Strides Pharma Science^TM^ received marketing authorization in November 2018. RAS coverage increased steadily after its introduction in April 2019, and a high RAS coverage was reached after 3-4 months (Fig 6c). Neither the timing of RAS distribution and trainings, nor other individual factors and contextual circumstances explained the observed patterns of RAS administration (data not shown).

RAS was distributed to health workers at the community level immediately following their initial trainings. A total of 126,904 RAS units were procured for the three countries, and 91,189 units were distributed to over 8,000 trained CHWs and primary healthcare workers. The remaining RAS units were either distributed after the implementation period, stored for future distribution, donated, or retrieved and incinerated if the recommended shelf-life had been reached. By 2020, the proportion of trained CHWs and peripheral healthcare workers who were actively engaged in administering RAS and submitting monthly reports ranged from 92% to 100% in the project countries.

#### DRC

Distribution of RAS was from the provincial level to the Health Zones, and then to secondary health facilities. These facilities then distributed RAS to the primary health facilities and CHWs. The nurses in the secondary health facilities were responsible for monitoring commodity availability and replenishing stock on a monthly basis. Four units of RAS were provided to each health care provider. Initially, CHWs and primary health care facilities had to submit requests for stock replenishment, but because of the high frequency of stockouts, this method was replaced by the supervisor bringing the RAS during each monthly supervision. According to the HCPS at midline and endline, over 80% of CHWs and PHCs surveyed had received at least one supervision visit during the past 6 months.

#### Nigeria

Distribution of RAS started at the State level, then to hospitals and further to the primary care/CHW level. Supervision visits and refresher trainings were conducted by different Federal and State actors, e.g. the National Malaria Control Programme/Federal Ministry of Health, Adamawa State Ministry of Health, and the Adamawa State Primary Health Care Development Agency. Hospital staff were responsible for supervising CHWs on a monthly basis. CHWs had to contact their supervisors when they were low on RAS. In practice, the CHW were then often asked to come to the health facility to pick new stock at their own expense. CHWs received only one pack (i.e. two capsules) of RAS at a time. The HCPS midline survey showed that 89.3% of CHWs and 97% of PHCs surveyed reported having received a supervisory visit within the last 6 months.

A particular issue in Nigeria was the very high temperature (over 35°C daily average) during the months of January to June. Although the capsule structure prevented the artesunate suppositories from leaking even at high temperatures, the absence of cooling devices at CHW and PHC levels reduced the shelf-life of RAS. Ongoing stability tests are being conducted and will be reported elsewhere (MMV manuscript in preparation).

A lockdown due to the COVID-19 pandemic was issued in Nigeria on the 30^th^ of March 2020 restricting the movement of people and supplies. This has undoubtedly contributed to the drop in RAS administration and PSS enrolments during spring 2020 (Fig 6b).

#### Uganda

Quarterly review meetings at PHC facility level were set up to track commodities for reporting to the national level. Since this system was not responding fast enough, a monthly district-level (parish) coordinator meeting was set up by CHAI and implemented to support overcoming logistics challenges. Supervision of CHWs was conducted in parallel with monthly parish coordinator meetings. Initially, CHWs were provided with one pack of RAS per quarter, based on predicted numbers of severe cases. This quantity was quickly shown to be insufficient, and subsequently CHWs were given at least two packs per quarter. According to the HCPS midline and endline surveys, over 85% of CHWs and 96% PHC facilities reported receiving at least one supervisory visit during the last 6 months. COVID-19 had also an impact on the movement of persons and on the supply chain from April to June 2020. In Fig 6c, we observed a slight decline in RAS administration over that period, as well as a stagnation in PSS enrolments.

### Behaviour Change Communication

In addition to supplying RAS, CARAMAL partners helped also to design, pre-test, and roll-out behaviour change communication (BCC) messages and materials in all three countries, to stimulate community level sensitization and demand on the introduction of RAS.

### RAS dosing

Evidence on dosing with RAS of study children was collected from slightly over 1000 study children (Fig 7). According to the manufacturer’s recommendations, children between 6 months and 3 years should receive one 100 mg suppository, while children from 3 to 6 years of age should receive two suppositories. This recommendations could be leading to variable dosing because of low-weight-for-age children. In the CARAMAL study, the recommended dosing was not universally adhered to, and many children were under-dosed. In DRC, 83% of children over 3 years that were administered RAS received one suppository instead of two. In Nigeria the situation was best with only 32% of children over 3 years receiving one suppository, while in Uganda this percentage was 55%.

**Fig 7:**
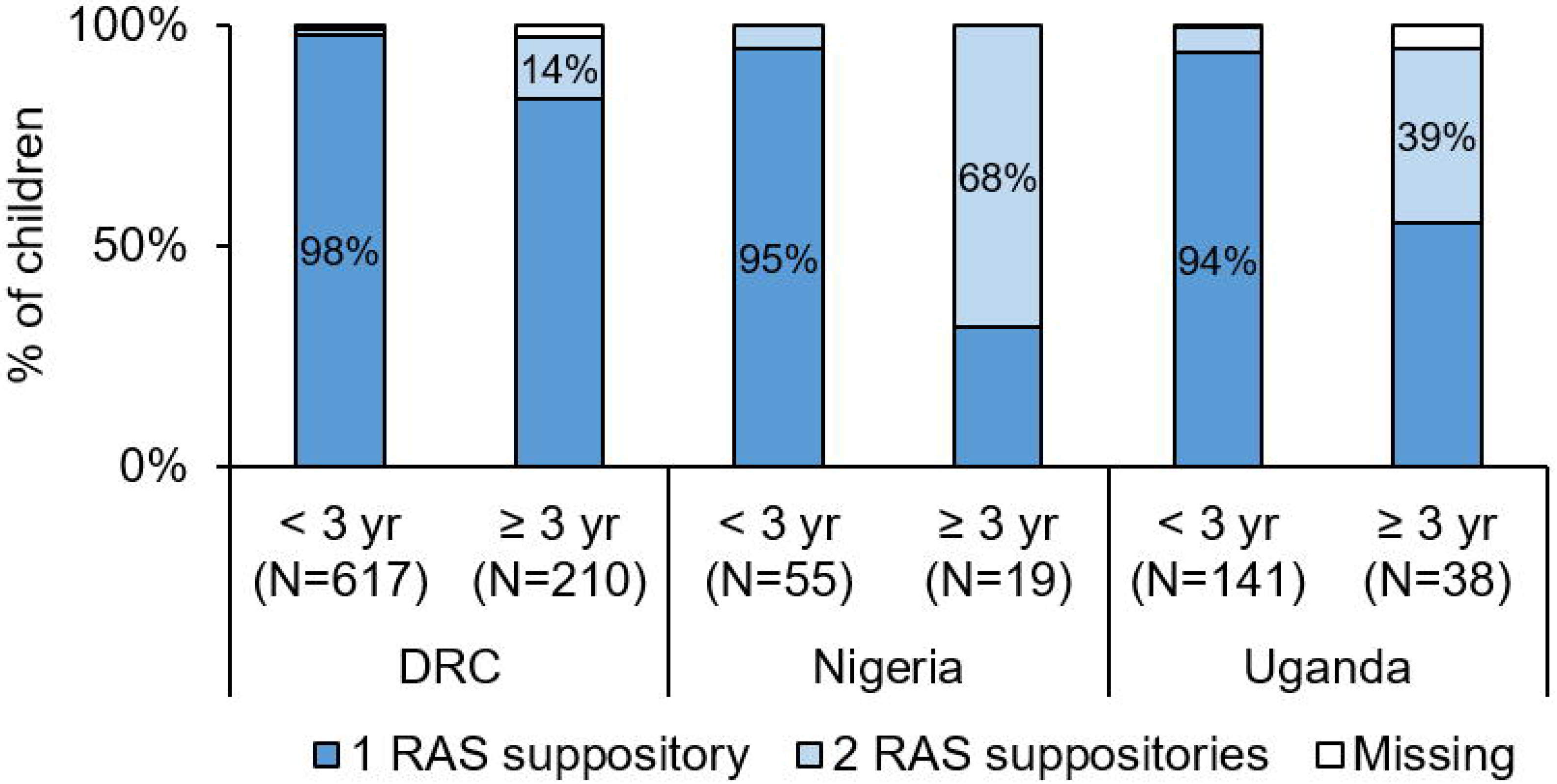
Number of suppositories received by enrolled children, by age group and country. yr=year. Missing=missing data.

### Key operational learnings

Key operational learnings from RAS implementation in the three project countries included:

1. Lower-than-expected coverage (DRC) and utilization (Nigeria, Uganda) of CHWs in the project areas, in part because of frequent commodity stock-outs and personnel fluctuations. This undoubtedly reduced confidence in the CHWs’ ability to provide adequate and reliable treatment.
2. Regular supportive supervisions substantially improved CHW engagement and performance, and were critical to a functioning RAS supply line. However, carrying out regular supervision visits was challenging from an operational, logistical and financial perspective – despite the fact that in all three settings this was a routine task expected to be performed by the health system.
3. Need for regular restocking of RAS at the CHW level and mechanisms for internal redistribution, given the high variability in RAS needs: some CHW regularly used RAS, while others did not use a single dose during the study period.
4. Identification of numerous barriers to referral completion [26], especially socio-economic factors
5. Need for additional investment in the quality of care provided for severe malaria treatment at referral health facilities, including the provision of a full course of ACT post-parenteral treatment to complete the treatment (Signorell et al. manuscript in preparation).
6. Need for the strengthening of routine data systems at the CHW level, including the procurement and supply management (PSM) systems, and formal/informal private care providers.

### Estimated rates of severe febrile illness in study children

It has been known for some time that there are many epidemiological, health system and clinical determinants driving the rates of severe febrile illness/malaria in a given setting [3]. Unfortunately, our empiric understanding of these determinants does not seem to have evolved much since the 1990s. Currently, rates of severe disease are derived entirely from modelling approaches [4] and reliable empirical estimates would usefully complement these approaches.

A rough estimation of the rates of severe febrile illness (independently of RAS treatment) could be computed within the CARAMAL study on the basis of the number of children with severe febrile illness recruited in the three sites (Table 2), and given the estimated number of children of that age group in the three settings (Table 1). DRC = 5,540 cases /145,107 children U5 = 0.038 or 38 per 1,000 for the 27 months of the study period; or *16.9 per 1,000 and per year*. **Nigeria**: 1,505/130,430 = 0.012 or 12 per 1,000 for the study period; or *5.3 per 1,000 and per year*. **Uganda**: 6,713/200,518 = 0.033 or 33.5 per 1,000 for the study period; or *14.9 per 1,000 per year*. These estimates are only approximations with substantial margins of error for at least four reasons. Firstly, the actual number of cases of febrile illness seen through the PSS is certainly below the real number occurring in the community, since not all children were brought to public providers for treatment. Unfortunately, we had no way to estimate this independently in this large population. Secondly, varying case definitions among the three sites undermined the representativeness of the number of reported cases of “severe febrile illness” (see also Table 3 below). Severe febrile illness episodes as defined by iCCM can have multiple causes (not only malaria), and represent on average a milder disease form than severe malaria as defined by the WHO (see below). Thirdly, access to prompt and effective antimalarial treatment at primary level is likely to influence the rate of progression from mild to severe disease [6, 13, 34]. Fourthly, up-to-date and accurate census figures were not available for the study areas, and hence the denominators were only an estimation based on best available data from public sources.

**Table 3:** Case definitions of severe febrile illness (as defined in the frame of iCCM) compared to the full medical definition of severe malaria.

| Severe febrile illness (iCCM [27]) | Severe *P. falciparum* malaria - WHO case definition. For laboratory values, see [10] |
| --- | --- |
| Fever over 37.5° C or a history of recent fever (essentially within the last 24 hours); plus one of the following danger signs:<br>– Lethargy/unconsciousness<br>– Refusal to feed/breastfeed<br>– Convulsions<br>– Repeated vomiting<br>In the DRC only:<br>– Unable to sit or stand up<br>mRDT for malaria is optional | One or more of the following, occurring in the absence of an identified alternative cause, and in the presence of *P. falciparum* asexual parasitaemia and in most cases fever.<br>– Impaired consciousness: A Glasgow coma score < 11 in adults or a Blantyre coma score < 3 in children<br>– Prostration: Generalized weakness so that the person is unable to sit, stand or walk without assistance<br>– Multiple convulsions: more than two episodes within 24 h<br>– Acidosis; severe acidosis manifested clinically as respiratory distress (rapid, deep, laboured breathing)<br>– Hypoglycaemia<br>– Severe malarial anaemia<br>– Renal impairment<br>– Pulmonary oedema<br>– Significant bleeding<br>– Shock<br>– Hyperparasitaemia |

All other parameters being equal, truly severe febrile episode case numbers were likely to be underestimated in Nigeria (low numbers of enrolled children with probably higher average severity in patients leading to high CFR (Fig 8) and overestimated in Uganda (low CFR indicative of low average severity in patients), while the DRC estimates were in-between. Nevertheless, these community severe febrile illness rates are probably among the best produced for any African setting to-date, and should have some utility.

**Fig 8:**
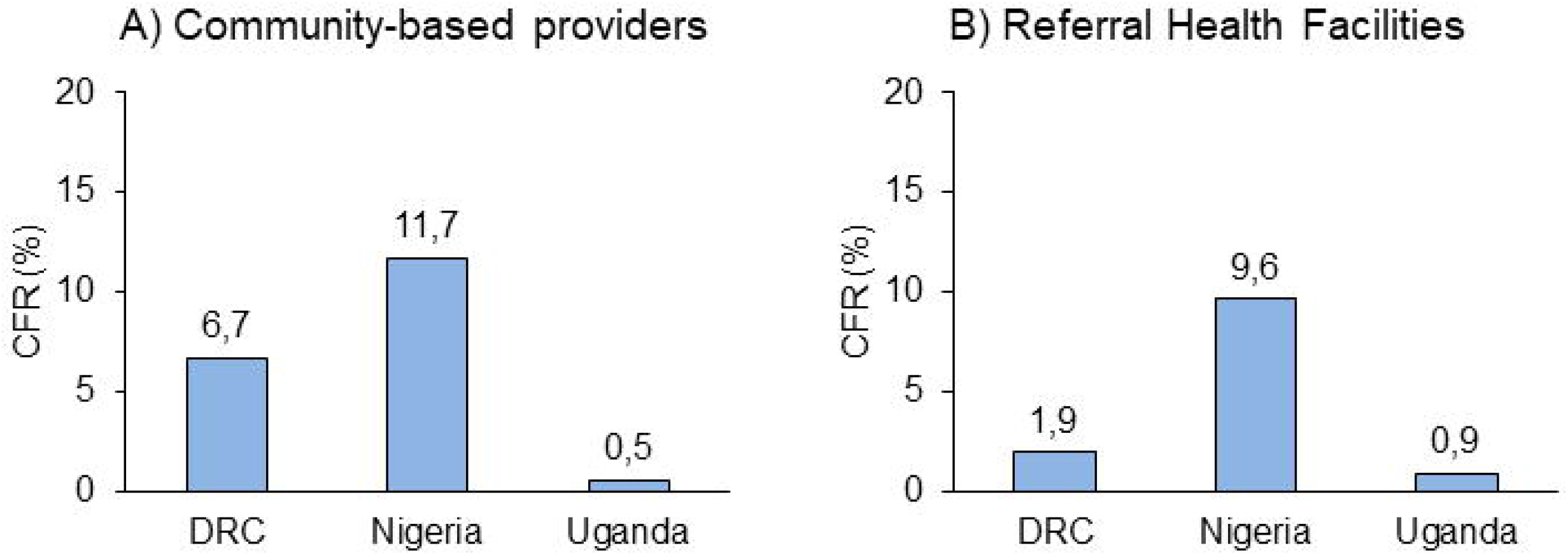
Overall case fatality ratio (CFR) across the entire study period, by enrolment location and country.

### Overall Case Fatality Rates (CFR)

The CFR differed significantly between the three countries, and in DRC also between both enrolment groups (Fig 8). The highest CFR were measured in Nigeria: 12.0% [95% CI 9.7-14.8] at community level and 10.0% [95% CI 8.2-12.3] at RHF level. The lowest CFR were found in Uganda (0.6% [95% CI 0.4-0.9] at community level and 0.9% [95% CI 0.6-1.3] at RHF level. In DRC, the CFR was substantially higher in community enrolments: 7.3% [95% CI 6.3-8.5], than in RHF enrolments: 1.9% [95% CI 1.4-2.6]. A detailed analysis of the CFR, a pre-post RAS impact analysis, as well as an individual risk factor analysis for mortality and severe disease are presented in a forthcoming publication [25].

### Case definitions: iCCM versus WHO definition of severe malaria

The difference in case definitions between severe febrile illness under iCCM and the classic WHO definition of severe malaria are shown in Table 3. A small but potentially significant difference between the two guidelines is that the target age group is different: below 5 years for iCCM, below 6 years in the WHO RAS guidelines. We did not assess the effect of this difference in the CARAMAL study.

This lack of overlap between the two case definitions has at least two implications for practice. Firstly, the inability to estimate correctly the true rates of severe malaria in the community has already been noted above, and represents an important limitation for tracking progress in malaria disease control. Possibly, recommending systematically an mRDT to febrile children with signs/symptoms of disease severity, could prevent unnecessary RAS pre-referral treatment. This approach has been shown to be feasible and safe [35]. This was also the case in the CARAMAL study.

Secondly, there seem to be a case for exploring in more detail the significance of some of the iCCM severity signs in children for their prognostic value, in a way similar to what was done for clinical and laboratory parameters of severe malaria [7]. This could for example be assessed for “Refusal to feed/breastfeed” or “Repeated vomiting”, which are not strictly indicators of severe disease. This could lead to re-assessing their value for initiating systematically a referral, with all its difficulties and uncertainties, and possibly recommend a wait-and-see strategy instead, with or without RAS administration.

## Conclusions

With over 9,000 episodes of severe febrile illness enrolled at the primary care level, the CARAMAL project aimed to document comprehensively key determinants of RAS effectiveness. The present project overview presented the project scope, objectives, methodological approaches, RAS implementation, and gave some key operational results. Using a comprehensive treatment seeking framework for severe febrile illness/severe malaria, the CARAMAL project documented systematically patients, communities and health systems in three remote, highly malaria-endemic setting (in DRC, Nigeria and Uganda). We also documented the RAS under-dosing problem, as well as the difficulties of getting enough RAS doses to all relevant health care workers in time. Finally, the comprehensive Patient Surveillance System was used to calculate empirical rates of severe febrile illness, as well as compute fatality rates of these patients, which were in excess of published values for hospitalized patients in two of the three countries (DRC and Nigeria).

Effective case management for severe malaria episodes requires a large number of actions to be completed in succession, including the availability of at least five commodities: an mRDT and RAS at primary level, a confirmatory test for malaria at the RHF, injectable artesunate (or any other approved parenteral drug) plus the required supportive treatments, and finally a full course of an ACT. This continuum of care was comprehensively documented by the CARAMAL project. As noted repeatedly elsewhere, improving the functioning of health facilities at all levels, as well as strengthening other sources of care (for example in the private sector), is key to lasting improvements of health in Africa, including for malaria [36–38]. A specific issue raised in the CARAMAL study was the non-overlapping case definitions between severe febrile illness and severe malaria, and we highlighted two practical issues arising from this.

A number of companion papers will present specific results on RAS impact [25], the referral process and its problems [26], post-referral care [Signorell *et al.* manuscript in preparation] and the cost-effectiveness of RAS [Lambiris *et al.*, submitted]. In addition, a number of country-specific publications will document in more detail the specifics of RAS implementation and impact in each project country.

The CARAMAL consortium’s experiences in rolling out RAS in the project countries generated a comprehensive understanding of specific operational challenges that other countries will confront, as well as potential solutions for overcoming them. Hence, it is hoped that CARAMAL will make a lasting contribution to improving malaria disease and mortality in the most highly malaria-endemic countries in the world.

## Data Availability

All study data used for producing the present manuscript are available from the corresponding author to individuals with a motivated request.

## Acknowledgements

We thank all the children and their caregivers who agreed to participate in this study; the health workers and local and national health authorities who provided their support. Individuals who contributed to this work are listed under Supporting information S1 Table. The authors acknowledge Silvia Schwarte from the Global Malari Programme at WHO for her support in the frame of the Enabler grant provided by UNITAID to WHO.

## Supporting information captions

S1 Table: Authors included under the group name “CARAMAL consortium”.

S1 Fig: Detailed study sites maps. Democratic Republic of Congo (a), Nigeria (b) and Uganda (c).

S2 Table: Key indicators and survey instruments used in the CARAMAL study, by research question.

S3 Table: Demographic characteristics of study population enrolled in the Patient Surveillance System (PSS), by enrolment location.

