## Supporting information for "Community access to rectal artesunate for malaria (CARAMAL): a large-scale observational implementation study in the Democratic Republic of the Congo, Nigeria and Uganda"

### Supplementary materials

#### S1 Table: Authors included under the group name "CARAMAL consortium".

| Name | Title | Affiliation |
|---|---|---|
| Marek Kwiatkowski | PhD | Swiss TPH, Basel, Switzerland |
| Nadja Cereghetti | MSc | Swiss TPH, Basel, Switzerland |
| Aurelio Di Pasquale | PhD | Swiss TPH, Basel, Switzerland |
| Robert Canavan | MA | Swiss TPH, Basel, Switzerland |
| James Okuma | PhD | Swiss TPH, Basel, Switzerland |
| Silvia Schwarte | MSc | World Health Organization, Switzerland |
| Hans Rietveld | BA | Medicines for Malaria Venture, Switzerland |
| Lydia Kabamba | Dr. | UNICEF Democratic Republic of Congo |
| Francine Kimanuka | Dr. | UNICEF Democratic Republic of Congo |
| Sylvie Luketa | Dr. | UNICEF Democratic Republic of Congo |
| Moulaye Sangare | Dr. | UNICEF Democratic Republic of Congo |
| Tony Byamungu | Dr. | UNICEF Democratic Republic of Congo |
| Emmanuel Emedo | Dr. | UNICEF Nigeria |
| Peter Baffoe | Dr. | UNICEF Nigeria |
| Oluseyi Olosunde | Dr. | UNICEF Nigeria |
| Halima Abdu | Dr. | UNICEF Nigeria |
| Linda Nsahtime-Akondeng | Dr. | UNICEF Nigeria |
| Garba Safiyanu | Dr. | UNICEF Nigeria |
| Flavia Mpanga | Dr. | UNICEF Uganda |
| Fred Kagwire | Dr. | UNICEF Uganda |
| Michael Mugwanya Musiitwa | Dr. | UNICEF Uganda |
| Joe Collins Opio | Dr. | UNICEF Uganda |
| Anita Mbadiwe | | CHAI Nigeria |
| Tayo Olaleye | Dr. | CHAI Nigeria |
| Remilekun Peregrino | | CHAI Nigeria |
| Abraham Okita | Dr. | CHAI Nigeria |
| Omowunmi Omoniwa | Dr. | CHAI Nigeria |
| Nnenna Ogbulafor | Dr. | CHAI Nigeria |
| Oluseyi Omokore | MD | CHAI Nigeria |
| Chizoba Fashanu | Dr. | CHAI Nigeria |
| Stella N. Nwosu | MD | CHAI Nigeria |
| Carine Olinga | | CHAI Democratic Republic of Congo |
| Jenny Bokanga | | CHAI Democratic Republic of Congo |
| Eric Mukomena | PhD | PNLP Democratic Republic of Congo |
| Jean-Claude Tembele | Dr | CHAI Democratic Republic of Congo |
| Albert Kadjunga | Dr | CHAI Democratic Republic of Congo |
| Fatou Mwaluke | MPH | CHAI Democratic Republic of Congo |
| Alex Ogwal | | CHAI Democratic Republic of Congo |
| Juliet Nakiganda | | CHAI Democratic Republic of Congo |
| Isaac Okiring | | CHAI Uganda |
| Maureen Amutuhaire | | CHAI Uganda |

**S2 Fig: Detailed study sites maps. Democratic Republic of Congo (a), Nigeria (b) and Uganda (c).**

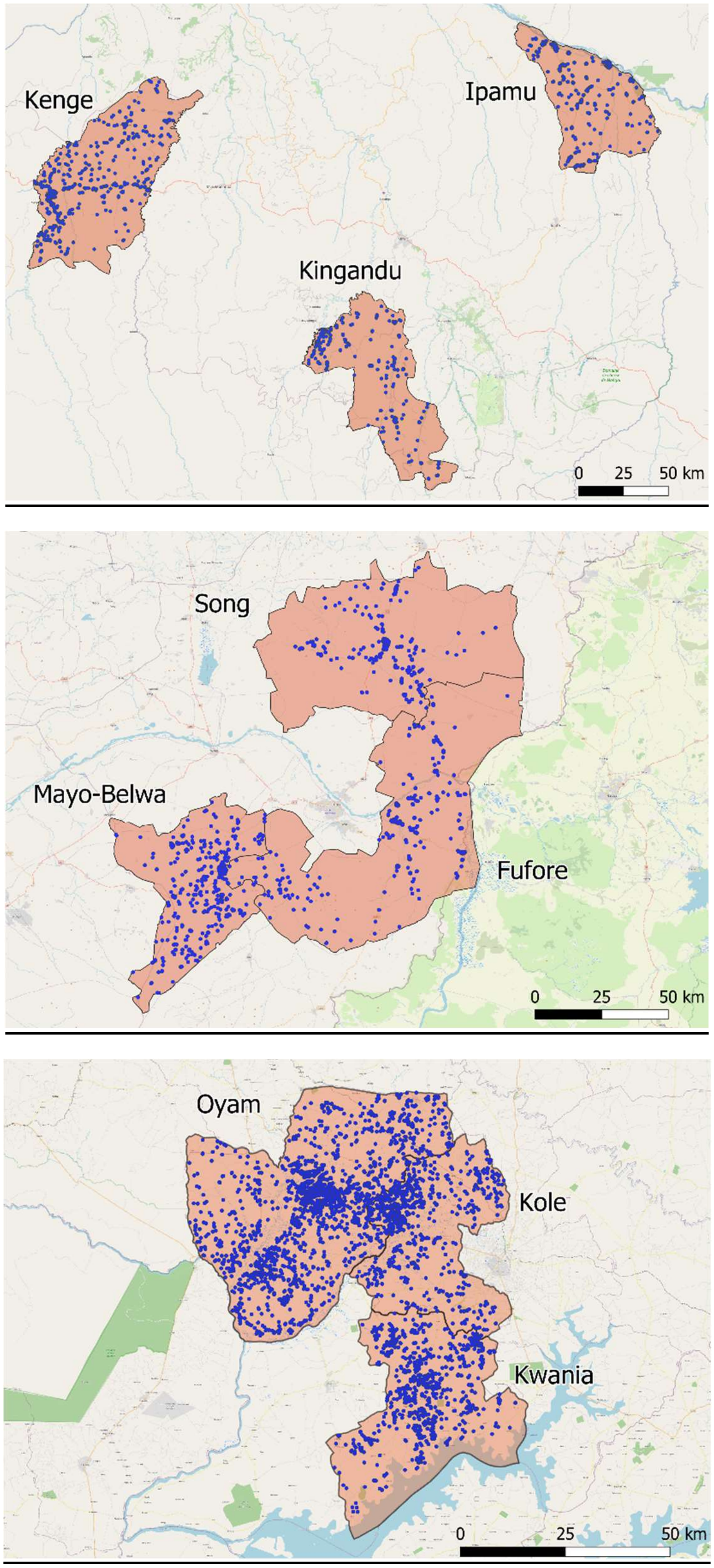

**S2 Table: Key indicators and survey instruments used in the CARAMAL study, by research question.**

PSS = Patient surveillance system; HCPS = Health care provider survey; HHS = Household survey; EES = Economic evaluation study; HF = Health facility; M&E = routine M & E activity.

| Research questions | Indicator | Source | Indicator description | Frequency |
|---|---|---|---|---|
| **I** | QA RAS coverage | M&E, HCPS | Proportion of trained and functional CHW and primary HF who provide QA RAS | Annually |
| | RAS availability | M&E, HCPS | Proportion of CHW/primary HF with RAS in stock | Annually |
| | Functional referral facilities | HCPS | Proportion of referral health facilities that have the capacity to manage severe malaria in children in line with global guidance | Annually |
| | Supervision | M&E, HCPS | Proportion of CHW/primary HF that received at least one supervisory visit in the past 3 months | Annually |
| **I + II** | Adverse events | PSS | Frequency of passively reported adverse events after RAS administration | Continuous |
| | Delayed haemolytic anaemia | PSS | Frequency of delayed haemolytic anaemia within 28 days after RAS administration | Continuous |
| **II** | Treatment seeking | HHS | Proportion of children <5 years with a recent history of fever (mild or severe) who attend CHW/primary HF | Annually |
| | CHW attendance | PSS | Proportion of children <5 years with a recent history of fever (mild or severe) who attend CHW/primary HF | Annually |
| | RAS acceptability among health workers | HCPS | Acceptability of pre-referral RAS among health workers | Annually |
| | RAS acceptability in community | HHS | Acceptability of pre-referral RAS among caretakers | Annually |
| | Referral | PSS | Proportion of children <5 years with severe febrile illness seen by CHW/primary HF who completed referral | Continuous |
| | Direct reporting | PSS | Number of children <5 years with severe | Continuous |

| | | | | |
|---|---|---|---|---|
| | | | febrile illness who report directly to RHF | |
| **I + III** | Pre-referral treatment | PSS | Proportion of children <5 years with severe febrile illness managed pre-referral according to guidelines | Continuous |
| | Post-referral treatment | PSS | Proportion of children <5 years with severe febrile illness managed post-referral according to guidelines | Continuous |
| **IV** | Case fatality ratio | PSS | Proportion of children <5 years with severe febrile illness seen by CHW/primary HF that resulted in death within 28 days | Continuous |
| | Malaria infection on day 28 | PSS | Proportion of children <5 years with severe febrile illness without parasites on day 28 | Continuous |
| | Successful referral | PSS | Proportion of children <5 years with severe febrile illness seen by CHW/primary HF who completed referral to a referral health facility | Continuous |
| **V** | Costs | EES | Total financial cost of managing severe case at community level. Marginal financial cost of adding RAS to the management. | Continuous From project records |
| | Cost-effectiveness | PSS EES | Financial cost of RAS interventions per death averted. | PSS and project records |
| **Exploratory / contextual** | Danger signs | HHS | Proportion of children <5 years with a recent history of fever who reported danger signs (signs of 'severe febrile illness'). | Annually |
| | Malaria prevalence | HHS | Proportion of children <5 years infected with malaria parasites | Annually |

**S3 Table: Demographic characteristics of study population enrolled in the Patient Surveillance System (PSS), by enrolment location.**
CHW=Community Health Worker. PHC=Primary Health Care (facility). RHF=Referral Health Facility.

| | Enrolment CHW/PHC | | Enrolment RHF | |
|---|---|---|---|---|
| | n | % | n | % |
| DR Congo | 3'042 | | 2'498 | |
| Health Zone | | | | |
| Ipamu | 1'128 | 37.1 | 1'163 | 46.6 |
| Kenge | 1'101 | 36.2 | 906 | 36.3 |
| Kingandu | 813 | 26.7 | 429 | 17.2 |
| Sex | | | | |
| Male | 1'615 | 53.1 | 1'315 | 52.6 |
| Female | 1'427 | 46.9 | 1'183 | 47.4 |
| Age (years) | | | | |
| 0 - 0.9 | 624 | 20.5 | 507 | 20.3 |
| 1 - 1.9 | 888 | 29.2 | 733 | 29.3 |
| 2 - 2.9 | 647 | 21.3 | 495 | 19.8 |
| 3 - 3.9 | 467 | 15.4 | 417 | 16.7 |
| 4 - 4.9 | 416 | 13.7 | 46 | 13.9 |
| Nigeria | 658 | | 847 | |
| LGA | | | | |
| Fufore | 270 | 41.0 | 313 | 37.0 |
| Mayo Belwa | 271 | 41.2 | 230 | 27.2 |
| Song | 117 | 17.8 | 304 | 35.9 |
| Sex | | | | |
| Male | 401 | 60.9 | 471 | 55.6 |
| Female | 257 | 39.1 | 376 | 44.4 |
| Age (years) | | | | |
| 0 - 0.9 | 76 | 11.6 | 94 | 11.1 |
| 1 - 1.9 | 181 | 27.5 | 220 | 26.0 |
| 2 - 2.9 | 186 | 28.3 | 263 | 31.1 |
| 3 - 3.9 | 132 | 20.1 | 162 | 19.1 |
| 4 - 4.9 | 83 | 12.6 | 108 | 12.8 |
| Uganda | 3'927 | | 2'786 | |
| District | | | | |
| Kole | 1'787 | 45.5 | 656 | 23.6 |
| Kwania | 1'055 | 26.9 | 580 | 20.8 |
| Oyam | 1'085 | 27.6 | 1'550 | 55.6 |
| Sex | | | | |
| Male | 2'093 | 53.3 | 1'553 | 55.7 |
| Female | 1'834 | 46.7 | 1'233 | 44.3 |
| Age (years) | | | | |
| 0 - 0.9 | 711 | 18.1 | 536 | 19.2 |
| 1 - 1.9 | 1'135 | 28.9 | 830 | 29.8 |
| 2 - 2.9 | 913 | 23.3 | 698 | 25.1 |
| 3 - 3.9 | 729 | 18.6 | 428 | 15.4 |
| 4 - 4.9 | 439 | 11.2 | 294 | 10.6 |
